# Beyond adherence: Experiences shaping engagement with oral anticancer medication among immigrant patients with haematological malignancies and limited dominant-language proficiency

**DOI:** 10.64898/2026.08.18.26360752

**Authors:** Sandra Michiels, Nathalie Meuleman, Sandra Tricas-Sauras

## Abstract

**Background:** Immigrant patients with limited dominant-language proficiency may face intersecting challenges when navigating cancer care and long-term oral anticancer treatment. Although studies have reported lower medication adherence among migrant and ethnic minority populations, less is known about how migration-related, linguistic, experiential and contextual factors shape treatment engagement from patients’ own perspectives. This study explored how immigrant patients experience illness, navigate treatment and engage with oral anticancer medication within the broader context of cancer care.

**Methods:** Thirteen immigrant patients with limited dominant-language proficiency receiving oral anticancer medication for haematological malignancies were recruited from the haematology outpatient clinic of a Belgian university hospital. Semi-structured interviews were conducted in participants’ native languages using an adapted version of the McGill Illness Narrative Interview, with professional interpreters or intercultural mediators. Interviews were analysed using inductive reflexive thematic analysis within an interpretivist framework.

**Results:** Analysis of patients’ illness narratives generated five experiential dimensions: 1) bodily, biographical and identity rupture; 2) temporal disruption and uncertainty; 3) linguistic vulnerability shaping the illness experience; 4) meaning-making and explanatory frameworks; and 5) resources sustaining treatment engagement. Linguistic vulnerability shaped access to biomedical knowledge, participation in healthcare encounters and patient autonomy, while patients mobilised personal, relational, existential, linguistic and institutional resources to sustain treatment continuity. Treatment engagement emerged as a dynamic and relational process embedded within broader migration-related, linguistic and healthcare contexts. Rather than representing fixed determinants or sequential stages, the five dimensions formed an evolving configuration whose relative salience varied throughout the illness trajectory.

**Conclusions:** This study proposes a multidimensional interpretive model of engagement with oral anticancer medication among immigrant patients with limited dominant-language proficiency. Rather than conceptualising adherence as an isolated individual behaviour, the findings show how migration-related contexts shape the conditions under which treatment engagement becomes possible, difficult or fragile. By foregrounding immigrant patients’ lived experiences, the study identifies experiential, linguistic, relational and structural dimensions of cancer care that are difficult to capture through behavioural adherence measures alone and offers insights for more equitable, context-sensitive and patient-centred oncology care.

## INTRODUCTION

Haematological malignancies are among the most common cancers worldwide, with an estimated 1.3 million new diagnoses and nearly 700,000 deaths each year(1). Their global burden is expected to rise substantially over the coming decades(2). The growing use of oral anticancer medications (OAMs)(3) has transformed treatment by shifting care from hospital-based administration to self-management at home. While this transition has improved convenience and quality of life for many patients, it has also transferred much of the responsibility for treatment from healthcare professionals to patients, making long-term treatment engagement and medication adherence major clinical challenges. In haematology, non-adherence is associated with preventable hospitalisations, disease progression, and increased mortality(4). Yet reliable data on adherence to OAMs remain scarce and are largely limited to chronic myeloid leukaemia (CML) and acute lymphoblastic leukaemia (ALL)(5) (6).

Despite growing interest in adherence to OAMs, oncology research—and particularly onco-haematology—has largely prioritised treatment outcomes and adherence measures over understanding cancer care from patients’ perspectives and lived experience. Consequently, little is known about how patients understand their illness, negotiate uncertainty, experience oral cancer treatment in everyday life, and sustain engagement with treatment over time.

In Belgium, adherence to OAMs in haematological malignancies has been insufficiently studied. The only available national data, limited to CML patients treated with imatinib, revealed alarmingly low adherence rates(7,8). More broadly, Belgian adherence research has focused on chronic conditions such as diabetes(9,10), heart failure(11) or HIV(12,13) without examining the role of migration background. This gap is particularly relevant because the Brussels Capital Region is one of the most linguistically and culturally diverse urban areas in Europe, where nearly half of residents were born abroad and more than one hundred languages are spoken(14). Although Belgium provides universal health coverage and intercultural mediation services, migrants and ethnic minorities continue to face linguistic, administrative, and health-literacy barriers that contribute to inequalities in access to care and health outcomes(15).

International studies consistently show that migrants and ethnic minorities are at greater risk of non-adherence across a range of chronic conditions(16,17). In cancer, minority patients appear less likely to adhere to treatment, potentially contributing to disparities in survival(18–20). However, most evidence originates from English-speaking high-income countries and rarely concerns haematological malignancies. In Belgium, no studies have examined the relationship between migration background and adherence to OAMs. Beyond documenting disparities, existing studies rarely examine the mechanisms through which they emerge or how they are shaped by migration-related experience and contexts, sociocultural conditions, and interactions with healthcare systems. This limitation echoes a broader evolution in migrant health research, which increasingly argues that migration should not be treated as an explanatory characteristic in itself but rather as a context in which multiple structural, linguistic, relational, and organisational factors interact to shape patients’ health experiences, treatment engagement, and ultimately treatment outcomes(21). This perspective provides an important conceptual framework for understanding the mechanisms shaping patients’ engagement with treatment among immigrant patients receiving OAMs.

To address this gap, we conducted a mixed-methods study comprising a prospective questionnaire-based survey followed by the present qualitative investigation. The quantitative component identified disparities in medication adherence and treatment-related experiences between immigrant and native-born patients. However, it could not explain how these disparities arose within patients’ lived experiences or how migration-related contexts shaped treatment engagement, providing the rationale for the present qualitative study.

To explore these dimensions, we conducted semi-structured interviews with thirteen immigrant patients using the McGill Illness Narrative Interview (MINI)(22), a framework designed to explore how individuals make sense of illness through explanatory models, prototypes, chain-complexes, and illness narratives. To better capture issues relevant to immigrant patients with (LDLP), we added two sections addressing language barriers and migratory experiences to the interview guide.

This qualitative study aims to explore how immigrant patients with LDLP experience, understand and navigate OAMs in Belgium by situating medication-taking within broader migration-related, biographical, and healthcare experiences. Rather than seeking to explain treatment engagement through migration itself, the study examines how migration-related contexts shape the conditions under which treatment engagement becomes possible, difficult, or fragile. By examining engagement with oral anticancer medication through immigrant patients’ perspectives and lived experiences, the study seeks to identify the experiential dimensions through which treatment engagement is constructed and to provide a conceptual framework that helps explain disparities in medication adherence observed among immigrant patients with haematological malignancies. In doing so, it develops a dynamic multidimensional interpretive model of engagement with OAMs and illustrates how patients’ perspectives can enrich understanding of treatment engagement beyond behavioural adherence measures alone.

## METHODS

### Participants, recruitment and terminology

This qualitative component constituted the qualitative strand of a broader mixed-methods study and adopted an interpretive, narrative-based design grounded in the MINI(22). Narrative approaches recognise that meaning is co-constructed through interaction and shaped by the contexts in which accounts are produced. Accordingly, the study relied on a semi-structured, flexible, and reflexive design that allowed participants to articulate their experiences in their own terms while ensuring comparability across interviews.

The study adhered to established standards for qualitative health research (COREQ, SRQR)(23). Participants were recruited prospectively from the haematology outpatient clinic of a Brussels university hospital (OECI-accredited comprehensive cancer centre) between 21^th^ of June 2021 and 8^th^ of September 2022. Recruitment aimed to capture a diverse range of clinical, linguistic, and migratory profiles. For the purposes of this study, the term ‘immigrant’ was used as an operational descriptor referring to foreign-born individuals, consistent with Belgian and European epidemiological conventions.

Eligible participants were adults (≥18 years) with a confirmed haematological malignancy, receiving at least one OAM for a minimum of one month, and presenting LDLP, defined as a partial or complete inability to communicate independently in French or Dutch during healthcare encounters, requiring linguistic mediation to ensure adequate understanding of study procedures and interview content. Patients with acute, uncontrolled psychiatric conditions were excluded.

Screening and enrolment were conducted by a practitioner-researcher with access to consultation schedules and electronic medical records. Information sessions were conducted in participants’ mother languages with professional interpreter or intercultural mediator support. Written informed consent was obtained from all participants, including consent for the use of their anonymised data in scientific and medical publications.

Before the interview, participants completed the same sociodemographic, linguistic, clinical, and self-reported measures used in the quantitative strand of the MADESIO mixed-methods study. These included age, gender, country of birth, length of residence in Belgium, language proficiency, and clinical characteristics, including haematological diagnosis. Information on usual communication arrangements during haematology consultations, including the involvement of relatives or linguistic mediators, was also collected. This ensured consistency across study components and enabled characterization of the clinical, linguistic, and migratory diversity of the qualitative sample. Because the qualitative strand relied on interpreter support and was not restricted to the six translated languages available in the quantitative study, it also enabled inclusion of patients with more severe language barriers and more recent migration trajectories. Detailed descriptions of the questionnaires and procedures have been published and are available(24).

### Sample size

The sample size of thirteen participants was determined pragmatically, in line with the interpretive aims of the study and the depth of narrative material generated through the interviews. In keeping with the epistemological assumptions of reflexive thematic analysis, the study did not seek theoretical saturation. Instead, the analytic objective was to develop a rich and conceptually coherent thematic interpretation grounded in the diversity of participants’ narrative.

### Interview procedure: adapted MINI

Interviews followed the French version of the MINI (22), a semi-structured narrative interview developed by D. Groleau and colleagues at McGill University to explore how individuals make sense of illness through narratives, explanatory models, prototypes, and illness trajectories. The MINI has been widely used in qualitative and transcultural health research to investigate illness experience across diverse cultural contexts.

The interview comprised five sections exploring 1) illness onset and early interpretations; 2) illness prototypes; 3) explanatory models; 4) treatment and healthcare trajectories; and 5) the personal, relational, and existential impact of illness. To better capture experiences particularly relevant to immigrant patients with LDLP and haematological malignancies, two additional sections were added: 6) the impact of language barriers on illness experience and treatment, and 7) cultural background as a potential resource – or non-resource – in coping with illness.

Consistent with the MINI’s philosophy, interviews were conducted in a semi-structured manner. The interview guide provided structure and comparability across interviews while allowing participants to elaborate, clarify, and expand upon their narratives.

### Multilingual data collection, translation and linguistic mediation

All interviews were conducted in the participants’ native languages with the assistance of professional interpreters or intercultural mediators. Professional interpretation was preferred to family interpretation in order to ensure confidentiality, minimise protective filtering of information, and preserve the integrity of participants’ narratives.

Interviews were audio-recorded and conducted using consecutive interpretation. Interpreters occasionally reformulated questions, clarified ambiguities, or adapted phrasing to participants’ communicative styles. Rather than being treated as methodological deviations, these interventions were understood as part of the co-constructed nature of qualitative interviewing.

All interviews were transcribed verbatim in the source language by native-speaking professional transcribers and subsequently translated into French by independent professional translators. The researcher reviewed the source-language transcripts alongside the original recordings before translation to identify omissions or inconsistencies, and the French translations afterwards to clarify ambiguities and discuss any remaining issues with translators when necessary.

Because the researcher did not speak all native languages, these procedures were not intended to independently verify linguistic or conceptual accuracy. Rather, they were designed to enhance transparency, traceability, and quality control throughout the production of the multilingual dataset.

Consistent with recommendations for multilingual qualitative research, analysis relied on professionally translated transcripts rather than on the interpreter’s live rendering during the interview. Translation was understood as an interpretive, not merely a technical one. Consequently, translated transcripts were treated as interpretive artefacts, and attention was paid throughout the study to the linguistic mediations through which meaning was produced.

Interpreters were briefed regarding confidentiality, fidelity, and consistency. Rather than assuming interpreter neutrality, the study recognised linguistic mediation as part of the co-construction of qualitative data and documented its influence through reflexive memoing and analytic discussions.

### Analytical approach

Data were analysed using Reflexive Thematic Analysis (RTA) as developed by Braun and Clarke(25), following their six recursive phases. The analysis was grounded in an interpretivist epistemology and conducted on the French translated transcripts, which constituted the unified analytic corpus.

Coding proceeded inductively, first on a patient-by-patient and section-by-section basis, following the structure of the adapted MINI. The interview sections served as a heuristic framework for comparing narratives across participants while codes remained grounded in participants’ accounts. Subsequent theme development involved cross-case comparisons and the identification of patterns of meaning that transcended individual interviews and MINI sections.

Themes were developed on the basis of conceptual coherence rather than frequency. A cross-sectional matrix was developed to identify patterns across participants and interview domains. Interpreter-mediated interactions were examined reflexively by comparing the live French rendering provided during interviews with the subsequent French translation of the source-language transcripts. Differences such as summarisation, omissions, reformulations, or shifts in emphasis were documented in analytic memos.

The final thematic structure emerged through iterative movement between coding, memo writing, cross-case comparison, and repeated engagement with transcripts and audio recordings. Analysis was conducted manually, and a detailed audit trail documenting coding decisions, theme development, reflexive notes, and analytic matrices was maintained throughout the study.

### Researcher reflexivity

The researcher occupied a dual position as practitioner-researcher within the haematology department. This positionality was integrated as a methodological resource throughout the study. Following each interview, reflexive notes documented contextual elements, interactional dynamics, positionality effects, and situations in which clinical or institutional knowledge may have shaped the exchange. These notes were revisited during coding and theme development.

Reflexivity also extended to the triadic configuration of interpreter-mediated interviews. Interpreter contributions were considered part of the interpretive context through which narratives were produced rather than treated solely as sources of bias. Analytical anticipations linked to clinical experience or to knowledge generated through the quantitative strand were explicitly documented and bracketed through memoing and repeated examination of the data.

### Ethical considerations

The study was conducted in accordance with the Declaration of Helsinki and received approval from the institutional Ethics Committee (BECT B0792021000018; internal reference CE3422).

Participants received detailed information regarding study objectives, voluntary participation, confidentiality, and their right to withdraw at any time. Particular attention was paid to ensuring informed consent among participants with LDLP through translated materials and/or professional interpreter support. The researcher’s independence from the clinical team was explicitly clarified in order to minimise any perception of therapeutic obligation.

All personal identifiers were removed from transcripts and quotations. Hospitals, clinicians, and locations mentioned by participants were replaced with neutral codes to preserve confidentiality while maintaining narrative coherence. Participant characteristics are reported in aggregate to minimise the risk of re-identification through combinations of indirect identifiers. Participants were also offered the opportunity to receive a summary of the study findings.

### Use of artificial intelligence tools and technologies

Generative artificial intelligence (ChatGPT) was used during the preparation of Fig 1 to support the visual design and graphical organisation of the conceptual model of engagement with oral anticancer medication (OAMs). The underlying conceptual framework, interpretation of the findings, and synthesis of the qualitative data were developed by the authors. All AI-generated suggestions were critically reviewed, modified where appropriate, and validated by the authors, who take full responsibility for the final content of the figure and manuscript.

**Fig 1.**
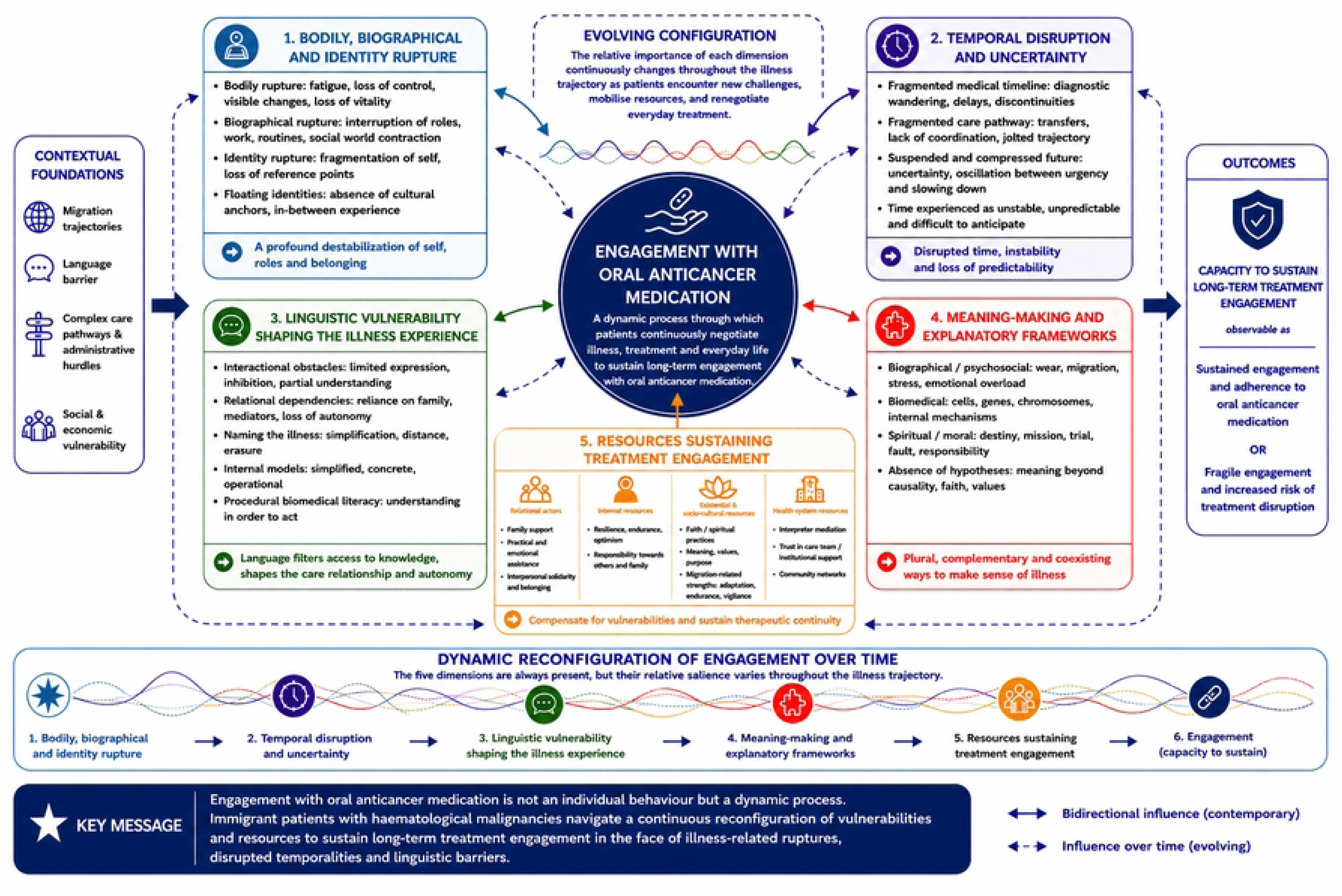
Multidimensional model of oral anticancer medication engagement among immigrant patients with haematological malignancies.

## RESULTS

Our inductive reflexive thematic analysis resulted in a multidimensional conceptual model of engagement with OAMs among immigrant patients with haematological malignancies and limited dominant-language proficiency. The analysis identified five conceptually coherent experiential dimensions that together provide an interpretive understanding of how engagement with OAMs is experienced, negotiated, and sustained throughout the illness trajectory: illness-related bodily, biographical and identity rupture; temporal disruption and uncertainty; linguistic vulnerability; meaning-making and explanatory frameworks; and relational and structural resources sustaining treatment engagement. Rather than representing fixed determinants or sequential stages, these dimensions constitute an evolving configuration whose relative salience changes throughout patients’ illness trajectories. Fig 1 illustrates the conceptual model and the dynamic reconfiguration of these experiential dimensions over the illness trajectory.

The model integrates five interconnected experiential dimensions whose relative salience evolves throughout the illness trajectory. Contextual foundations shape their dynamic interplay, while resources may compensate for vulnerabilities and sustain therapeutic continuity. Solid bidirectional arrows represent contemporary reciprocal influences; dashed arrows represent influences evolving over time.

### Participant characteristics

Thirteen patients participated in the study [Table 1]. Participants ranged in age from 20 to 76 years and represented heterogeneous clinical, linguistic, and migratory profiles. Migration trajectories ranged from recent arrival in Belgium (<1 year for the most recently arrived participant) to more than 25 years of residence, with participants differing in country of birth, haematological disorder, and self-reported and clinician-assessed French proficiency. Table 1 also summarises usual communication arrangements during routine haematology consultations and the linguistic and methodological characteristics of the qualitative interviews. Interviews were conducted in Brazilian and European Portuguese, English, Polish, Moroccan Arabic, Romanian, Russian and Turkish, illustrating the multilingual context in which the study was conducted. Interview duration ranged from 41 to 111 minutes.

**Table 1.** Sociodemographic, clinical, linguistic and interview characteristics of the qualitative study sample.

| Characteristic | Category | n |
| --- | --- | --- |
| **Sociodemographic and migratory characteristics** |  |  |
| Gender | Women | 9 |
|  | Men | 4 |
| Age group (years) | 20–40 | 4 |
|  | 41–60 | 6 |
|  | 61–74 | 2 |
|  | ≥75 | 1 |
| Country of birth | Brazil | 3 |
|  | Morocco | 2 |
|  | Poland | 3 |
|  | Portugal | 1 |
|  | Romania | 2 |
|  | Türkiye | 1 |
|  | Ukraine | 1 |
| Length of residence in Belgium | <3 years | 2 |
|  | 3–10 years | 1 |
|  | 10–20 years | 7 |
|  | >20 years | 3 |
| **Clinical characteristics** |  |  |
| Haematological disorder | Acute myeloid leukaemias and related precursors | 1 |
|  | Chronic myeloid neoplasms | 5 |
|  | Mature B-cell disorders | 1 |
|  | Myeloproliferative neoplasms | 2 |
|  | Plasma cell neoplasms | 4 |
| **Linguistic and communication characteristics** |  |  |
| Self-reported French proficiency | Not at all | 0 |
|  | Very little | 6 |
|  | Moderately | 7 |
|  | Fluently | 0 |
| Haematologist-assessed French proficiency | Not at all | 3 |
|  | Very little | 3 |
|  | Moderately | 6 |
|  | Fluently | 0 |
|  | Not available | 1 |
| Relative present during routine consultations | Yes | 7 |
|  | No | 6 |
| Usual communication arrangement | Relative mediation | 6 |
|  | Professional interpreter | 1 |
|  | English | 2 |
|  | None | 4 |
| **Qualitative interview characteristics** |  |  |
| Interview language | Brazilian Portuguese | 3 |
|  | English | 1 |
|  | Moroccan Arabic | 2 |
|  | Polish | 2 |
|  | European Portuguese | 1 |
|  | Romanian | 2 |
|  | Russian | 1 |
|  | Turkish | 1 |
| Interpreter profile during qualitative interview | Intercultural mediator | 1 |
|  | Social interpreter | 7 |
|  | Professional interpreter | 5 |
| Interview duration | ≤69 min | 7 |
|  | 70–89 min | 2 |
|  | ≥90 min | 4 |

**Note.** French proficiency was assessed both by participants and by their treating haematologist. One participant was interviewed over two separate sessions.

The thematic dimensions presented below should not be interpreted as independent categories or successive stages of the illness trajectory. Rather, they represent complementary dimensions through which participants described their experiences of illness, treatment, and care. Although presented separately for clarity, they form an evolving experiential configuration whose relative salience varied across participants and over time. Together, these dimensions provide an interpretive understanding of how treatment engagement is experienced, negotiated, and sustained throughout the illness trajectory.

### Theme 1 — Bodily, biographical and identity rupture

#### Bodily rupture: weakening and loss of control

Patients described illness as profoundly disrupting their relationship with their body. Fatigue, weakness, and reduced physical capacity interrupted everyday routines and undermined previously taken-for-granted forms of autonomy. As one participant explained, “Before, I did everything by myself… now I can no longer do anything properly” (P2). For many, bodily limitations imposed new rhythms and dependencies, reshaping daily life and future possibilities. Visible bodily changes, including weight loss and altered appearance, further reinforced this disruption. Beyond symptoms, participants described a sense of dispossession and loss of continuity with their former selves.

#### Biographical rupture: disruption of social roles and contraction of the social world

Illness also disrupted the social and biographical structures that had previously organised daily life. Participants described being forced to stop working, abandon routines, and reduce social activities. Work, often associated with dignity, purpose, and stability, was experienced as a particularly significant loss. As Patient 4 stated, “I am sad to stop working.” Beyond their practical consequences, these disruptions affected family roles, self-esteem, and social participation. Illness imposed a reorganisation of priorities and everyday responsibilities, reshaping patients’ relationships to time, work, and social presence.

#### Identity rupture: fragilisation of the self and loss of reference points

Beyond bodily and biographical changes, illness challenged participants’ sense of identity. Several described feeling emotionally fragile or no longer recognising themselves. As Patient 13 explained, “My personality has changed… I used to be strong; now I feel weak.” Cognitive difficulties, including memory lapses and reduced concentration, further destabilised internal reference points and contributed to feelings of discontinuity. For some, this identity disruption occurred within broader migratory trajectories already marked by unstable belonging and weakened cultural reference points. Participants often described being the first or only person in their family to experience cancer and reported few cultural narratives through which to interpret the illness. In this context, illness emerged as an experience without familiar scripts or shared points of reference.

Taken together, these findings illustrate how haematological illness constituted a profound bodily, biographical, and identity-related rupture. For many participants, this disruption resonated with earlier experiences of migration, displacement, and social reorganisation, contributing to a cumulative sense of instability across multiple domains of life.

### Theme 2 — Temporal disruption and uncertainty

Haematological cancer profoundly disrupted participants’ experience of time, creating a rupture between a “before” and an “after,” fragmented care trajectories, and a future that felt uncertain, suspended, or compressed.

#### Fragmented trajectories and disrupted continuity

Participants frequently described prolonged periods of uncertainty before diagnosis. Symptoms were initially minimised, self-managed, or attributed to benign causes. As Patient 2 explained, “I didn’t go… I had no pain… I was just tired, weak,” while Patient 3 managed symptoms for months by repeatedly taking pain medication. For others, delays resulted from repeated consultations, inconclusive investigations, or difficulties accessing specialist care. Patient 7 recalled that despite persistent exhaustion, her physician “didn’t send me for tests.”

Temporal fragmentation often continued after diagnosis. Participants described trajectories marked by transfers between hospitals, successive referrals, delays, and partial explanations. As Patient 12 explained, “I was hospitalized in Hospital A… they took me to Hospital B… then I went to Hospital C.” These discontinuities made it difficult to understand how care was organised or to anticipate what would happen next. Time was experienced as disjointed, unpredictable, and largely beyond personal control.

#### A suspended future: between urgency and slowing down

Illness introduced profound uncertainty regarding prognosis, treatment duration, and future possibilities. Participants frequently questioned how long the illness would last or whether recovery was possible. As Patient 8 asked, “How long will this last?… Will it ever end?” This uncertainty narrowed temporal horizons and made future projection difficult. Some participants described a collapse of futurity, expressing feelings that the future had become fragile, opaque, or inaccessible.

In response, two contrasting temporal orientations emerged. For some, awareness of mortality generated urgency and a desire to act while time remained available. Patient 12 explained, “Now I am in a hurry. I want to do things quickly… to fulfil my dreams.” Others described illness as an invitation to slow down, reassess priorities, and focus on self-care. As Patient 10 reflected, “It made me slow down… my priorities changed.”

These orientations were not mutually exclusive and often coexisted within the same narrative. Participants oscillated between planning and waiting, acceleration and pause, hope and uncertainty. Medical appointments, examinations, and treatments’ imposed rhythms that frequently conflicted with personal temporalities, reinforcing the impression of a future that remained suspended and difficult to grasp.

Taken together, these findings illustrate how haematological illness profoundly reshaped participants’ relationships to time. Fragmented care trajectories, diagnostic uncertainty, and an unstable future contributed to experiences of temporal disorientation that complicated participants’ ability to anticipate, plan, and make sense of their illness trajectories. For many participants, these experiences unfolded within broader migration-related trajectories marked by administrative uncertainty, disrupted continuity, and ongoing adaptation, further shaping how illness and treatment were experienced over time.

### Theme 3 — Linguistic vulnerability shaping the illness experience

Language emerged as a central dimension shaping access to biomedical knowledge, therapeutic relationships, patient autonomy, and engagement with treatment.

#### Language, relational dependence, and restricted participation

Many participants described language barriers as a source of inhibition, uncertainty, and reduced participation during consultations. Although some understood parts of medical discussions, they often felt unable to ask questions, clarify information, or engage in deeper conversations with clinicians. As Patient 9 described, there was “a kind of blockage” that prevented her from seeking clarification. Others contrasted these experiences with consultations conducted in their native language, where they felt able to negotiate, express opinions, and participate more actively in decisions.

Language barriers also generated forms of relational dependence. Participants frequently relied on spouses, relatives, interpreters, or bilingual acquaintances to communicate with healthcare professionals, arrange appointments, or navigate administrative procedures. As Patient 1 explained, “When I thought they might not understand, I waited for my husband to come.” The descriptive characteristics of the participants further supported this observation: among those who routinely attended haematology consultations with a companion, communication was most often facilitated by that relative, whereas only one participant reported relying on a professional interpreter during routine consultations. These findings suggest that family members frequently assumed an essential role in mediating communication, extending their contribution beyond emotional support to include linguistic and organisational functions. Such dependencies reduced patients’ autonomy and increased vulnerability in everyday healthcare interactions.

Beyond family-mediated communication, routine haematology consultations were characterised by diverse communication arrangements. While several participants relied on relatives to facilitate communication, others communicated primarily in English or attended consultations without any identified linguistic support. Although the present study cannot determine whether these different arrangements adequately met patients’ communication needs, they illustrate the diversity of communication arrangements observed during routine haematology consultations. Communication therefore relied on diverse formal and informal resources, whose mobilisation appeared to vary across participants and clinical encounters.

Despite these constraints, participants developed compensatory strategies to maintain agency. These included using translation applications, searching for information online, preparing consultations in advance, relying on written materials, and requesting interpreter support when available.

#### Understanding illness: from naming to procedural literacy

Participants developed diverse ways of understanding their illness despite limited access to biomedical terminology. Naming practices ranged from generic expressions (“my illness”, “a blood disease”) to simplified biomedical categories such as “blood cancer” or “bone cancer.” For some, the name of the illness remained emotionally difficult to pronounce or was deliberately avoided in order to protect themselves or their families. These practices reflected varying degrees of appropriation, distance, and emotional engagement with the diagnosis.

Beyond naming, many participants relied on simplified internal models to make sense of complex haematological conditions. Illness was translated into familiar bodily categories such as “sick cells,” “rebellious cells,” or anaemia. These representations did not reflect ignorance but rather attempts to construct understandable explanations using accessible linguistic and cognitive resources. Such models provided interpretive anchors that enabled patients to orient themselves despite the complexity of biomedical discourse.

At the same time, many participants demonstrated a practical form of biomedical literacy focused on treatment management rather than disease mechanisms. They described treatment through routines, biological monitoring, medication cycles, and observable outcomes. As Patient 8 explained, treatment meant “three weeks every night, and then there is a one-week break.” Others relied on biological results to orient themselves and monitor disease progression: “When I get blood tests every three months… sometimes he tells me it’s stable, sometimes slightly lower” (P13). This procedural understanding enabled participants to follow treatment regimens, monitor changes, and integrate medication into everyday life.

Linguistic vulnerability shaped not only communication but also the forms of knowledge available to patients. Through naming practices, simplified explanatory models, and procedural understandings of treatment, participants developed workable forms of biomedical literacy that enabled them to engage with treatment despite persistent limitations in access to biomedical information.

### Theme 4 — Meaning-making and explanatory frameworks

Participants drew on multiple explanatory frameworks to make sense of the origins of their illness. These frameworks frequently coexisted and complemented one another rather than being mutually exclusive. Explanations could be biographical, psychosocial, biomedical, spiritual, or moral, while some participants mobilised no causal explanation at all. Contrary to common assumptions, cultural background was rarely invoked as a direct cause of illness. Instead, migration experiences, personal spirituality, and family values played a more prominent role in helping patients restore coherence, reduce uncertainty, and maintain a sense of agency in the face of illness. Explanatory frameworks therefore functioned not only as causal accounts but also as interpretive resources through which illness could be integrated into broader life narratives.

#### Biographical and psychosocial explanations

For many participants, illness was understood as the consequence of accumulated strain rooted in personal biographies, including hard work, migration-related difficulties, experiences of violence, loneliness, years of sacrifice, precarious living conditions, or psychological suffering. As Patient 9 explained, “I arrived in a foreign country… I couldn’t speak French… my family is in Poland… I am alone here.” These explanations situated illness within broader life trajectories and helped restore coherence to disruptive experiences.

#### Biomedical explanations

Some participants drew on biomedical explanations involving genes, chromosomes, cells, or internal bodily mechanisms. These explanations ranged from relatively precise references, such as chromosomal abnormalities, to broader notions of hereditary vulnerability or “sick cells.” Rather than replacing other explanatory frameworks, biomedical explanations typically coexisted with biographical, psychosocial, or spiritual interpretations, reflecting partial but meaningful appropriations of medical knowledge.

#### Spiritual and moral explanations

Several participants mobilised spiritual or moral frameworks that provided meaning rather than biomedical causality. Illness could be interpreted as destiny, a life mission, a personal trial, or a test of endurance. As one participant reflected, “It is a mission… I must go through this to improve” (P12). Others described forms of guilt or moral responsibility linked to past experiences. These frameworks provided coherence, direction, and emotional containment in situations characterised by uncertainty and limited control.

#### Meaning beyond causality

Not all participants sought causal explanations. Some explicitly stated that they had never considered why the illness occurred. In these cases, meaning emerged through non-causal sources such as faith, family values, or personal dispositions shaped by migration and life experience. Rather than explaining illness, these resources helped patients live with uncertainty and maintain continuity in everyday life.

Participants relied on diverse explanatory frameworks to make sense of illness. These frameworks frequently coexisted rather than competed, illustrating the plurality of ways through which immigrant patients interpreted illness and integrated it into their broader life experiences. Rather than providing definitive causal explanations, they offered interpretive resources that helped participants restore coherence and live with uncertainty.

### Theme 5 — Resources sustaining treatment engagement

Participants mobilised a wide range of personal, relational, existential, linguistic, and institutional resources that supported treatment engagement throughout the illness trajectory.

#### Everyday adaptation and treatment continuity

For many participants, medication intake became highly routinised and integrated into everyday life. Treatment was often described as habitual or automatic — “now it is a habit” (P4); “it’s like a cup of coffee: you get up and drink it” (P7). This consistency was supported by trust in physicians, fear of disease progression, and personal discipline shaped by demanding life trajectories. Yet participants also described occasional lapses related to fatigue, travel, or everyday pressures. As Patient 9 acknowledged, “Sometimes it happens that I don’t take it.” These experiences highlighted adherence as a dynamic process of routines, adjustments, interruptions, and recovery rather than a fixed behavioural state.

#### Relational and mediating resources

Family support emerged as a central pillar of treatment engagement. Relatives organised medication, accompanied patients to appointments, translated information, and provided emotional encouragement. For some, engagement depended directly on the vigilance of family members. As Patient 13 explained, “She stays with me until I take the treatment… she checks everything.”

Linguistic mediation constituted another important resource. Family members, interpreters, and bilingual acquaintances frequently acted as intermediaries between patients and healthcare professionals. While this support enabled access to care and information, it also reflected forms of dependency created by linguistic vulnerability. Treatment engagement therefore emerged as a collective rather than purely individual process.

#### Personal, existential and migratory resources

Participants also mobilised personal and existential resources to sustain treatment over time. Determination, endurance, and perseverance were frequently described as essential for coping with side effects, uncertainty, and prolonged treatment. As Patient 5 stated, “a lot of grit, a lot of determination… I will overcome all of this.” These dispositions were often rooted in previous life experiences, family values, and migration trajectories marked by adaptation and hardship. As one participant reflected, “Because I am an immigrant. When you live in a country that is not your own, you learn to be patient” (P13). Such experiences appeared to foster endurance, adaptability, and tolerance of uncertainty that participants later mobilised in response to illness.

Faith served as an important source of reassurance and strength for some participants, although others explicitly rejected spiritual interpretations. As Patient 5 explained, “Faith is a form of trust… it gives you strength.” Family-based values such as patience, responsibility, and perseverance also provided moral support. Importantly, these resources differed from the explanatory frameworks described in Theme 4: rather than explaining why illness occurred, they helped patients continue living with illness and maintain engagement with treatment despite uncertainty.

#### Institutional resources: safety, stability and gratitude toward Belgium

A recurring theme across narratives was a strong sense of gratitude toward Belgium and its healthcare system. Participants frequently contrasted their current situation with experiences of insecurity, instability, or limited healthcare access in their countries of origin. Access to treatment, financial protection, and continuity of care provided participants with a strong sense of security. As Patient 12 observed, “Here, you have everything… they give me all the medications”.

For many participants, treatment engagement was therefore sustained not only by personal effort and family support, but also by confidence in the institutions providing care.

Taken together, these findings illustrate the diversity of personal, relational, existential, linguistic, and institutional resources that participants mobilised to sustain treatment over time. Within migration-related contexts marked by illness, uncertainty, and linguistic challenges, these resources appeared to support continuity of treatment and everyday adaptation.

## DISCUSSION

This qualitative study provides the first in-depth exploration of how immigrant patients with LDLP living with haematological malignancies in Belgium experience illness, navigate oral anticancer medication, and sustain engagement with treatment. Drawing on immigrant patients’ perspectives captured through illness narratives, the study offers a qualitative interpretive model of engagement with oral anticancer medication, illustrating how migration-related, linguistic, temporal, and relational contexts shape patients’ experiences of treatment over time. In doing so, it contributes to a richer understanding of engagement by examining oral anticancer medication through the lens of patients’ lived experiences.

Our findings suggest that adherence is not simply an individual behaviour but is embedded within a multidimensional experiential process encompassing illness-related rupture, temporal disruption, linguistic vulnerability, meaning-making, and relational and structural resources. Together, these thematic dimensions provide an interpretive model of how immigrant patients experience and sustain engagement with oral anticancer medication over time. Rather than representing independent explanatory factors, these experiential dimensions constitute an evolving configuration whose relative salience is continuously reconfigured throughout the illness trajectory. At different moments, illness-related rupture, temporal disruption, linguistic vulnerability, meaning-making, and available resources assume different degrees of prominence. Engagement with oral anticancer medication therefore emerges from this evolving configuration rather than from the influence of any single dimension. This perspective extends existing work on chronic illness, migration, and medication adherence by showing how patients’ perspectives illuminate dimensions of engagement that are difficult to capture through behavioural adherence measures alone.

Importantly, these findings should not be interpreted as suggesting that migration itself explains treatment engagement. Rather, they indicate that migration-related contexts shape the conditions under which treatment engagement becomes possible, difficult, or fragile. This interpretation is consistent with recent developments in migrant health research, which increasingly move beyond essentialist understandings of migration towards contextual and structural explanations of health inequalities(21).

The adapted MINI proved particularly valuable in revealing forms of reasoning extending beyond causal explanatory models. It highlighted the relative absence of salient prototypes, which may reflect the rarity, heterogeneity, and limited public visibility of haematological malignancies rather than migration itself, as well as the prominence of explanatory frameworks, chain-complex reasoning, and existential resources that might have remained less visible using approaches focused exclusively on causal explanations. Together, these findings enrich our understanding of how patients organise uncertainty and sustain engagement with treatment.

A first thematic dimension illuminated by participants’ narratives is the biographical rupture experienced by LDPL immigrant patients, echoing Bury’s concept of biographical disruption(26) and Charmaz’s description of the loss of the familiar body(27). Participants described a collapse of bodily continuity, social roles, and identity, producing a sense of estrangement that was often amplified by migratory trajectories marked by precarity, instability, and the loss of familiar reference points.

These findings align with studies showing that haematological malignancies generate substantial physical and psychological burdens (28–36). Similar experiences of bodily and social disruption have also been described among migrants living with other chronic conditions, suggesting that migration-related precarity may amplify illness-related rupture(37,38).

In our study, bodily rupture often translated into identity rupture: the body ceased to support the self and instead became a source of limitation and uncertainty. This disruption was intensified by two features specific to the migratory context: the absence of comparable family or community narratives through which to interpret the illness, and a fluctuating or suspended sense of cultural belonging that could not readily be mobilised as a resource.

The Common-Sense Model of self-regulation(39) and the Necessity–Concerns Framework(40) provide a useful lens for understanding how identity disruption may affect treatment engagement by altering perceptions of treatment necessity and concerns. Although this mechanism was not directly assessed, these frameworks suggest a plausible pathway linking biographical disruption to adherence difficulties.

This interpretation is consistent with qualitative research showing that long-term require identity reconstruction(41,42) and that disruptions to bodily continuity, autonomy, and social roles can undermine patients’ capacity to sustain long-term adherence (43). Our findings extend this literature by showing how these processes unfold within migration, linguistic vulnerability and structural instability.

A second thematic dimension emerging from participants’ narratives concerns the profound temporal disorganisation produced by illness. Diagnostic delays, fragmented care pathways, unpredictable therapeutic trajectories, and difficulties projecting into the future disrupted patients’ experience of time. Such disruptions have been described in the chronic illness literature(44,45), but appear particularly acute in haematological malignancies, where disease trajectories are often unstable and unpredictable(46)(47).

Participants described prolonged periods of uncertainty before diagnosis, characterised by repeated consultations, referrals, and delayed explanations, echoing Walter et al.’s concept of patient delay(48). As Castañeda(49) and Willen(50) have argued, migration itself is often characterised by discontinuous temporalities; illness therefore becomes embedded within a broader “continuity of discontinuities.

Temporal instability persisted after diagnosis. Participants described fragmented care trajectories, disrupted continuity, and difficulties anticipating what would happen next. These experiences resonate with Hsueh et al.’s notion of reduced temporal agency(51), whereby patients lose the ability to anticipate, organise, and negotiate healthcare rhythms. This loss was further reinforced by limited health literacy, linguistic vulnerability, and social isolation, all of which complicated navigation of care and reduced opportunities to align personal temporalities with institutional schedules.

Paust’s concept of temporal capital(52) provides a useful lens for interpreting these findings. Temporal capital refers to the capacity to understand, anticipate, and negotiate the temporal demands of healthcare. In our study, this capacity appeared constrained by linguistic barriers, limited familiarity with the healthcare system, and reduced informational and relational resources, making it more difficult to anticipate appointments, understand care pathways, coordinate administrative procedures, and align personal with institutional schedules.

Participants also described profound uncertainty regarding prognosis, treatment duration, and future possibilities. Many struggled to imagine or plan for the future, reflecting what Clayton(53) and Mishel(54) describe as structural ambiguity. This contraction of the future has been widely described in oncology(55–57), where patients often oscillate between urgency and slowing down rather than experiencing these orientations as opposites.

Temporality emerged as a constitutive dimension of treatment engagement. Temporal disorganisation undermines patients’ ability to anticipate, plan, and sustain long-term therapeutic routines. Among immigrant patients with LDLP, these difficulties were further amplified by migration-related uncertainties, limited health literacy, and linguistic barriers, offering a second experiential process through which engagement with oral anticancer medication may become fragile.

The narrative analysis also brought to light a third experiential dynamic shaping treatment engagement: linguistic vulnerability as a structural determinant of the illness experience. While numerous studies have associated language barriers with lower medication adherence, poorer treatment understanding, reduced healthcare utilisation, and increased complications(58,59), our findings illuminate the mechanisms through which linguistic vulnerability operates in the context of malignant haematology.

Participants described a form of linguistic inhibition characterised by anticipatory stress, fear of speaking incorrectly, and difficulties expressing concerns or seeking clarification. This experience echoes Birkelund et al.’s notion of linguistic vulnerability(60), whereby patients may feel cognitively intact yet unable to communicate effectively. Similar findings have been reported in language-discordant healthcare encounters, where communication barriers restrict the expression of emotions, symptoms, and uncertainty(61,62). In malignant haematology, where treatment pathways are complex and decisions may need to be made rapidly, such inhibition may compromise patients’ ability to understand therapeutic priorities and warning signs(63).

Even when communication remained possible, participants frequently described a gap between what they understood and what they could express. This dissociation between passive comprehension and expressive capacity has been widely documented in health literacy research(64,65) and may limit patients’ ability to formulate questions, verify understanding, and participate actively in clinical exchanges. Previous studies in oncology similarly suggest that limited proficiency in the dominant language is associated with a more fragile understanding of treatment protocols, risks, and side effects(66,67).

Linguistic vulnerability also shaped access to biomedical knowledge. Participants frequently relied on relatives, interpreters, or digital tools to obtain information and navigate care, reflecting patterns described in previous research(68,69). Consistent with broader work on health literacy(16,70,71), these strategies helped compensate for communication barriers but did not necessarily provide a comprehensive understanding of disease and treatment. Instead, many developed simplified representations of their illness using generic labels or accessible bodily categories, reflecting forms of partial appropriation of biomedical knowledge (72–76), alongside a procedural form of biomedical literacy centred on treatment routines, biological monitoring, and observable outcomes as often seen in the scientific literature. This procedural understanding aligns with research showing that patients often know how to follow treatment without fully understanding the underlying interacting dimensions(73,77,78). Importantly, it also closely resembles what the MINI conceptualises as chain-complex reasoning, whereby knowledge is organised around routines, sequential links, and observable effects rather than explicit causal explanations or salient prototypes.

Although these strategies enabled participants to engage with treatment, they did not necessarily support informed decision-making or autonomous management of complications. As previous research has shown(79–81), procedural understanding may coexist with important gaps regarding treatment mechanisms, risks, and side effects. In malignant haematology, where therapeutic protocols are highly technical and often evolve over time, these limitations may create persistent zones of clinical vulnerability.

Linguistic vulnerability is not simply a communication barrier but a structural condition that reshapes the entire illness experience. It influences access to biomedical knowledge, participation in clinical encounters, decisional autonomy, treatment understanding, and the mobilisation of social support. By revealing these interrelated processes, our study extends previous research on language barriers and adherence and demonstrates how linguistic vulnerability may generate a cascade of effects that undermine long-term engagement with oral anticancer medication.

By revealing these interrelated processes, our study extends previous research on language barriers and adherence and demonstrates how linguistic vulnerability may generate a cascade of effects that undermine long-term engagement with oral anticancer medication.

The descriptive characteristics of the participants further support this interpretation. Self-rated and clinician-assessed French proficiency did not systematically coincide, and one participant who self-rated his French as “*very little*” later explained during the interview that he knew only the words “*merci*” and “*au revoir*” and was otherwise unable to communicate. These observations suggest that a single self-rated question may not adequately capture the communicative competencies required in complex healthcare encounters. Rather than relying solely on patients’ self-assessment of language proficiency, communication needs may be better appreciated through their ability to understand information, express concerns, ask questions, and participate in clinical interactions. This perspective reinforces the conceptual distinction between language proficiency and linguistic vulnerability, the latter reflecting a relational and contextual condition shaped by both individual communicative resources and the organisation of care.

Our narrative analysis revealed a fourth thematic dimension shaping treatment engagement: the construction of meaning in a context where few salient prototypes were available. While explanatory models theory emphasises multiple modes of reasoning – including causal explanations, prototypes, analogies, and associative chains(82,82,83) – participants rarely relied on prototype-based reasoning. This contrasts with findings from other chronic conditions(84–87) and may reflect the rarity, heterogeneity, and limited public visibility of haematological malignancies with few culturally shared reference points through which patients can interpret their illness(88–91), rather than reflecting a characteristic of migrant populations.

In the absence of such prototypes—a phenomenon explicitly described within the MINI framework(22) —participants relied more heavily on causal explanations. Consistent with Young’s observations(82,92), these explanations frequently drew on biographical, psychosocial, moral, spiritual, or migration-related experiences to restore coherence in the face of uncertainty. Illness was commonly attributed to accumulated stress, overwork, emotional suffering, or the hardships associated with migration. Similar patterns of interpretation have been documented among patients living with haematological malignancies(93).

These interpretative frameworks shape expectations about treatment(94) and can be understood as coherent attempts to restore control under uncertainty, consistent with the Common-Sense Model(39). Participants also mobilised dispositions shaped through migration – including patience, endurance, vigilance, and adaptability – to cope with illness. Similar findings have been reported among migrant populations living with chronic and infectious diseases(86,95–97). In our study, these migratory dispositions complemented these interpretative resources by helping patients confront uncertainty and sustain engagement with treatment despite limited biomedical understanding.

Previous research suggests that acknowledging patients’ explanatory models can strengthen mutual understanding and treatment engagement(98–100). Our findings extend this perspective by showing that when patients felt unable to share their interpretations, opportunities for dialogue and shared understanding became more limited.

Meaning-making played a central role in sustaining treatment engagement. In a context where salient prototypes were largely absent, patients drew on explanatory frameworks and migration-shaped dispositions to restore coherence, navigate uncertainty, and maintain engagement with illness and treatment.

Building on the previous thematic dimensions, treatment engagement emerged as a relational, moral, and organisational process requiring multiple resources to sustain therapeutic continuity over time. This interpretation aligns with contemporary psycho-oncology frameworks, which conceptualise coping as a multidimensional and evolving process mobilising emotional, social, practical, and existential resources simultaneously(101–104).

Participants relied on a range of personal, familial, and existential resources to sustain engagement with treatment. Consistent with previous research(105–108), resilience, perseverance, meaning-making, responsibility towards others, and migration-shaped dispositions supported continuity despite uncertainty, side effects, and prolonged treatment.

Theoretical work on translocational dispositions(109) and empirical studies of migrant health (110) suggest that experiences of precarity, adaptation, and institutional navigation may cultivate forms of endurance, vigilance, and flexibility that can later be mobilised in response to illness. Our findings support this perspective, showing that migration functioned not as a cultural explanatory framework but as a reservoir of practical and moral resources that helped patients confront uncertainty and remain engaged with treatment.

Therapeutic engagement also emerged as a form of distributed agency, shared across patients, relatives, interpreters, and healthcare professionals. This finding is consistent with research showing that adherence to oral anticancer medication is often co-produced within relational network(94) and that in Belgium, professional interpreters, although valued for their accuracy and neutrality, remain difficult to mobilise in routine care because of organisational constraints(111). Participants frequently relied on relatives or interpreters to access information, navigate care, and communicate with clinicians, illustrating how treatment engagement depended on resources extending beyond the individual patient. Our reflexive analysis further supports methodological work showing that interpreters are not neutral conduits of information but active participants in the co-construction of meaning(62,80).

Institutional resources also played a significant role. Consistent with previous studies(113–115), trust in healthcare professionals and confidence in the healthcare system emerged as important facilitators of treatment engagement. For participants whose migration trajectories had been marked by instability, precarity, or limited access to care, the Belgian healthcare system represented a source of continuity, security, and reassurance. This institutional anchoring appeared to reinforce engagement with treatment and strengthen confidence in long-term care.

By articulating five coexisting thematic dimensions within a single conceptual model, this study shows that engagement with oral anticancer medication cannot be solely understood through individual beliefs, behaviours, or treatment characteristics. Instead, migration-related vulnerabilities become translated into difficulties sustaining long-term treatment through a dynamic interplay of relational, linguistic, temporal, and structural processes.

More broadly, the findings support a shift from viewing adherence as an individual behaviour towards understanding it as a dynamic, relational, and context-dependent process embedded within broader social, linguistic, and institutional environments. The proposed model should therefore be understood as explanatory rather than predictive. Its purpose is not to quantify the relative influence of individual mechanisms on adherence, but to explain how these mechanisms are experienced, articulated, and interact within patients’ illness narratives.

### Clinical and policy implications

The multidimensional model developed in this study has several implications for the care of immigrant patients with LDLP living with haematological malignancies. First, the findings suggest that engagement with oral anticancer medication should not be approached solely as an individual behaviour but as a dynamic experiential process shaped by an evolving configuration of illness-related rupture, temporal disruption, linguistic vulnerability, meaning-making, and relational and structural resources. Clinicians should therefore attend not only to medication-taking behaviours but also to the broader conditions shaping engagement over time.

The findings highlight the importance of recognising linguistic vulnerability as a structural determinant of care rather than a simple communication barrier. Rather than relying solely on patients’ self-reported language proficiency, healthcare teams may need to assess communication needs throughout the care trajectory, taking into account patients’ ability to understand information, express concerns, ask questions, and participate in clinical interactions. Ensuring timely access to professional interpreters and adopting iterative, multimodal approaches to communication may help reduce the gap between procedural and conceptual understanding of treatment. Creating opportunities for patients to discuss their explanatory frameworks may further strengthen mutual understanding and reduce the risk of unrecognised misunderstandings. More broadly, communication support should be viewed as a flexible component of care rather than a one-size-fits-all intervention. While family members frequently contribute to everyday communication and care navigation, access to trained professional interpreters may be particularly important during clinically complex or high-stakes encounters, such as diagnosis disclosure, treatment initiation, or discussions of disease progression. Tailoring communication support to patients’ evolving needs may strengthen continuity of care while promoting meaningful participation in clinical decision-making.

At organisational and policy levels, the results underscore the importance of carefully addressing both linguistic vulnerability and temporal precarity as structural dimensions of inequality in cancer care. Although Belgium’s intercultural mediation program constitutes an important resource, participants’ narratives suggest that access remains inconsistent in practice. Strengthening access to professional interpretation services, simplifying administrative procedures, reducing navigational complexity, and providing multilingual resources that support both procedural and conceptual understanding may help reduce the vulnerabilities identified in this study. Policies that promote continuity of care, stable care pathways, and accessible communication support throughout the illness trajectory may be particularly important for patients whose migration trajectories have been marked by instability and uncertainty.

Consistent with the proposed conceptual model, supporting engagement with oral anticancer medication requires interventions that address not only patients’ individual capacities, but also the evolving linguistic, temporal, relational, and organisational contexts in which treatment is experienced.

### Limitations and future research

Several limitations should be considered when interpreting these findings. The study included thirteen immigrant patients with limited dominant-language proficiency receiving treatment for haematological malignancies in a single comprehensive cancer center in Brussels. While enabling an in-depth exploration of an underrepresented population in research, the way these experiential dimensions are expressed and configured may differ across healthcare systems, institutional settings, or linguistic environments.

The study focused specifically on foreign born immigrants with limited proficiency in the dominant language. Consequently, the experiential configuration described here may differ among immigrants who are fluent in the host-country language, second-generation populations, or other groups facing different forms of vulnerability. In addition, no native-born comparison group was included. This study was designed to generate an in-depth understanding of treatment engagement within a specific migratory and linguistic context rather than to attribute treatment engagement to migration itself or to determine whether the identified dynamics are unique to immigrant patients. Some of the experiential dimensions identified here may also be relevant to other patients living with complex chronic illnesses; however, the present study does not allow conclusions regarding their specificity or relative salience beyond the population studied.

Future research should examine the transferability of the proposed model across different migrant populations, clinical settings, and healthcare systems. Comparative studies may help clarify how these thematic dimensions are configured under different structural conditions and how they shape treatment engagement across diverse populations. Longitudinal qualitative studies could further explore how the relative salience of these experiential dimensions evolves throughout the illness trajectory and across key transitions in care. Further methodological work is also needed to refine approaches to multilingual qualitative research and to better understand how linguistic mediation shapes both healthcare encounters and the production of research data. Such work would contribute to refining and further testing the conceptual model proposed in the present study across diverse clinical, linguistic, and organisational contexts.

## CONCLUSION

This study provides the first qualitative exploration of engagement with oral anticancer medication among immigrant patients with LDLP living with haematological malignancies in Belgium. The findings suggest that engagement with oral anticancer medication is best understood as a dynamic experiential process shaped by an evolving configuration of illness-related rupture, temporal disruption, linguistic vulnerability, meaning-making processes, and relational and structural resources whose relative salience evolves throughout the illness trajectory.

Drawing on immigrant patients’ lived experiences and illness narratives, this study proposes a multidimensional interpretive model of engagement with OAMs that helps explain how migration-related contexts shape the conditions under which treatment engagement becomes possible, difficult, or fragile. Beyond adherence as a behavioural outcome, the findings highlight how migration-related vulnerabilities become translated into everyday challenges of understanding, navigating, and living with illness, thereby shaping patients’ capacity to sustain engagement with oral anticancer medication over time.

Promoting equitable engagement with OAMs therefore requires moving beyond individualistic models of adherence towards recognising the broader social, linguistic, relational, and institutional conditions through which treatment engagement is experienced and sustained. More broadly, these findings demonstrate how immigrant patients’ perspectives and lived experiences can enrich understanding of engagement with oral anticancer medication beyond behavioural adherence measures alone. By bringing patients’ perspectives into the study of treatment engagement, this work offers insights that may inform more equitable, context-sensitive, and patient-centred oncology care.

## Data Availability

The qualitative interview transcripts underlying this study cannot be made publicly available because they contain detailed personal, clinical, linguistic and migration-related narratives that may permit participant re-identification despite the removal of direct identifiers. Public sharing would also be inconsistent with the conditions of informed consent, which restricted the use and transfer of participants’ data to the purposes and persons specified in the approved study documentation. Relevant anonymised excerpts supporting the findings are included within the manuscript. Requests concerning access to additional data may be directed to the institutional Ethics Committee, subject to applicable ethical and data-protection requirements.

## ACKNOWLEDGMENT

We express our deepest gratitude to all the patients who generously shared their time, experiences, and trust. Their narratives form the heart of this study, and this work would not have been possible without their participation.

We warmly thank the intercultural mediators and professional interpreters whose expertise, availability, and commitment ensured the linguistic and cultural integrity of the interviews.

Their contribution was essential to conducting rigorous and ethically grounded multilingual research.

We are sincerely grateful to the authors of the McGill Illness Narrative Interview for granting permission to use the interview guide and for their valuable methodological guidance. Their work provided a conceptual and narrative framework that greatly enriched this study.

We also thank Mrs. Chbaklo Noura, Master’s student in Public Health, who conducted two of the thirteen interviews with exemplary rigor and professionalism as part of her thesis work.

Finally, we acknowledge the financial support of the Association Jules Bordet. Their contribution made it possible to secure the full set of resources required for interpretation, transcription, and translation, without which this multilingual qualitative study could not have been conducted.

